# Relationship between Dietary Patterns with Closure Growth Plate in 12-13 Years Old Girls in Kermanshah

**DOI:** 10.1101/2023.09.11.23292092

**Authors:** Mahsa mirian, Mohammadreza Rafiei, Tina Khosravy, Mehdi Moradinazar, Mehnoosh Samadi

## Abstract

**Background & Objective:** Bone linear growth during puberty depends on several factors, including nutrition. In addition, malnutrition in children and adolescents can affect both linear growth and growth plate chondrocytes. This study was conducted to determine the association between dietary patterns, bone age status, and closure growth plate in adolescent girls.

**Methods and Materials:** In this study, a validated semi-quantitative 167-items food frequency questionnaire was used to determine major dietary patterns. Major dietary patterns were also identified by the component analysis method. Furthermore, anthropometric data and information about physical activity were collected from participants.

**Results:** In the present study, 70.3% of participants had bone age less than chronological age, and the findings related to 3 main dietary patterns were identified (healthy diet, high salt and sugar diet, and western/mixed diets). Among the 3 main dietary patterns, a healthy dietary pattern showed a significant correlation with the difference between chronological age and bone age (B=-0.106). This research reveals that adherence to a healthy diet was associated with an increase in bone age (P=0.02).

**Conclusion:** The current study showed a significant association between healthy dietary patterns and the bone age of participants. Based on the results, it can be claimed that a lack of components in dietary patterns can have a negative effect on chondrocytes of growth plates. The present findings confirm that children and adolescents who followed suitable dietary patterns were less likely to be stunted.

## 1. Introduction

In recent years, nutrition has been described as one of the important factors in children’s and adolescents’ height. In all nutrition interventions, the theory is that growth and height will improve in children and adolescents with proper nutrition(Vidhyashree et al., 2015). One of the most important stages in life regarding healthy eating habits is adolescence. Studies show that eating behaviors in the adolescent years will also follow up during adulthood. Therefore, creating healthy eating behaviors in adolescents can significantly affect their health(Khajebishak et al., 2021). Unhealthy eating behaviors in the teenage years may result in a lack of essential nutrition that will finally affect their height growth(Nicklas et al., 2001). Change in body composition leads to increase growth rate and changes in puberty in general lead to increased nutrition needs of adolescents(Shekari et al., 2019). Puberty is a period of life that is accompanied by severe changes in height and body composition in such a way that during this period, height growth increases significantly. In this stage, men receive more muscular and skeletal mass, and women experience an increase in fat mass. Girls experiencing menstruation at a young age will face shorter height, more body mass index (BMI), and a risk for obesity(Jezewska-Zychowicz et al., 2017). In puberty, height growth in athletic children and adolescents can signify receiving enough energy for physical activity. Evaluating and measuring growth can be considered an index for general health or nutrition status in a population of children(Park et al., 2007).

The process of linear bone growth in growth plates happens due to chondrogenesis and a change in the shape of cartilage to the skeletal tissue(Esfarjani et al., 2013). Osteogenesis happens by distinguishing cartilage precursors mesenchymal cells to reproductive chondrocytes and adult chondrocytes that are metabolically active. During the reproduction and puberty of chondrocyte cells, the extracellular proteins that are the extracellular matrix components will be made by the endoplasmic reticulum, eventually leading to differentiation into osteoblasts(Whittle et al., 2012). Turning chondrocyte cells into adult cells stops their cell division, and their cell volume will be 5-10 folds. Then the chondrocytes are exposed to the planned cellular death(Hoppe et al., 2004). The cartilage will be replaced by the bones through extracellular matrix calcification and blood vessel destruction. Finally, it leads to the longitude growth of the bone(Hoppe et al., 2004; Joslowski et al., 2013; Muslimatun and Wiradnyani, 2016).

Vegetarian diets mostly focus on consuming vegetarian foods, including vegetables, fruits, beans, lentils, nuts, whole grains, and a small amount of fat. In such diets receiving food from animal sources like red meat, chicken, fish, dairy, and egg has been forbidden. Also, these diets do not include processed foods and sweets(Zárate-Ortiz et al., 2019). One of the significant efficiencies of the vegetarian diets index is giving positive points to vegetarian food and negative points to animal food. Generally speaking, it determines the quality of vegetarian diets is divided into 2 groups: healthy and unhealthy groups. It also differentiates between the whole vegetarian food and the processed ones(Baum et al., 2017). Vegetarian diets have beneficial effects on the incidence and course of different diseases such as obesity, diabetes type 2, cardiovascular diseases, cancer, and rheumatoid arthritis. The beneficial effects of vegetarian diets have significantly increased the number of people using this diet in recent decades. Also, the outcomes of the studies have shown that using an unhealthy plant-based diet index (uPDI) is along with an increase in the prevalence of short-stature in the first 4 months of children’s life. This influence has only been a result of unhealthy vegetarian diets, and plant-based diet index (PDI) and healthy plant-based diet index (hPDI) do not cause such effects(Melaku et al., 2018). In Asian communities, there is limited proof of vegetarian diets and chronic diseases(Fisher et al., 2002).

Regarding the classification of vegetarian diets, we will discuss the association between vegetarian diets index and bone growth in children based on un/healthy vegetarian diets, and the beneficial effects of vegetarian diets on various disorders and diseases. Also, the supplementation of essential nutrients, vitamins, and minerals needed for children’s growth through vegetarian diets, as nowadays malnutrition, an important, influential factor on children’s growth, is more prevalent. Healthy eating can play an important role in improving children’s physical condition and growth.

## 2. Materials and Methods

### 2.1. Research Method and the Participants

The current study was done in Kermanshah city in 2018. In this cross-sectional study, 350 girls in the age range of 12-13 were selected after examining their growth levels by endocrinologists and pediatricians have entered the study.

The sample size has been calculated by 90% power and a confidence coefficient of 95% in the current study. Because there’s no study to examine the relationship between vegetarian diet index and bone growth in adolescents, adult studies have been used to calculate the sample size(Ng, 2020). Considering 3 patterns, our sample size includes 216 participants. 20% has been added to the number of participants, and the sample size reached the number 350; this was done to increase the accuracy of the research on each nutrition pattern.

The criterion for entering the study includes filling out the consent to participate in the study and being a girl in the age range of 12-13 with open growth plates and normal height when they participate in the study. The exclusion criterion of this study included girls who have followed a diet (any diet) in the past 6 months, people with endocrine, cardiovascular, liver, kidneys, and thyroid diseases, and people who didn’t want to participate in the study for any reason. The criterion for entering the study and the method was explained completely in one meeting, and the written consent was received from the participants. This study has been confirmed by the Ethics Committee of Kermanshah University of Medical Sciences Research Council. The collected data include demographic information, physical activity level, food frequency questionnaire (FFQ), Anthropometric index, body composition, and the average bone age that we will discuss wholly in the following sections.

### 2.2. Anthropometric and Body Composition

The standing height of the participants was measured with no shoes on and while they were completely lying on the wall, and the SM 350 device with an accuracy of 0.5cm was used. The participants’ weight was measured using the Body Analyzer device In the Body 770 model. With the accuracy of 50 grams, it’s noteworthy that their weight was measured with no shoes on and while wearing light clothes(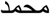 et al.). Other uses of the Body Analyzer device are measuring body composition, including BMI, percent body fat (PBF), soft lean mass (SLM), and Total Body Water (TBW). The collected data were used to calculate BMI, the outcome of the division of weight (kg) on the squared height (meters). The WHO standard BMI curves for girls (age range of 5-19 years old) were used to determine the participants’ BMI, and finally, their BMI centile for the age was found(Soliman et al., 2009). The international physical activity questionnaire evaluated the participants’ physical activity(Adriani and Wirjatmadi, 2014). This questionnaire has used some questions on children’s activity at school and out of it and the intensity of their physical activity during a week.

### 2.3. Determining the Openness of Skeletal Growth Plates and Evaluating Skeletal Age

To make sure the skeletal growth plates were open and to determine the skeletal age of the participants, dual-energy x-ray absorptiometry (DEXA) was used as a reference method to evaluate the body composition(Imdad and Bhutta, 2011). This method was performed by a radiologist on the dominant wrist of the participants.

### 2.4. Evaluating the Diet

The semi-quantity FFQ whose reliability and validity have been confirmed, was used to determine the participants’ dietary patterns. This questionnaire includes 167 foods that used the amount of intake nutrition by the participants in a year(Stammers et al., 2015; Min et al., 2015). Generally, vegetarian diets are divided into 3 groups:

1. PDI recommends using a variety of vegetarian food and reducing the amount of animal food.
2. hPDI recommends whole grains, fruits, vegetables, and nuts with beneficial effects.
3. uPDI focuses on using vegetarian food with less beneficial effects (purified grains, sweets, juices, and drinks sweetened with sugar).

These 3 patterns are related to the increased risk of diseases(Lee et al., 2012).

To determine the dietary pattern, 167 foods were classified into 22 classes (**Table 1**). This classification was done based on the similarity of the foods in the nutrients. If there are any differences in the food’s composition (like egg, fish, etc.), it would be placed individually in one group. The nutrition patterns were determined by the factor analysis method in which the varimax rotation is used to create a simple matrix and differentiator. The scree-plot test was used to determine the number of factors (dietary patterns)(Bhandari et al., 2001), and one point was based on the consumption charge of various nutrition groups for each dietary pattern, 1 point was assigned to each individual.

**Table 1.** Food groups used in the analysis of food patterns.

| **Food groups** | **food items** |
| --- | --- |
| Refined grains | White bread (loaves, baguettes), noodles, pasta, rice |
| Whole grains | Stone bread, Berber bread, corn, barley |
| beans | Lentils, beans, chickpeas, soybeans, other legumes |
| Red Meat | Beef, lamb, minced meat |
| Processed meats | Hamburger, sausage, sausage and pizza |
| Body meat | Heart, kidney, liver, tongue, brain, intestines |
| Chicken | Chicken |
| Fish | Types of fish |
| egg | egg |
| Low-fat dairy products | Low-fat milk, low-fat yogurt, buttermilk |
| Full fat dairy products | Full-fat milk, full-fat yogurt, cocoa milk, condensed yogurt, cream cheese, cream and milk, ice cream, regular cheese |
| the vegetables | Boiled potatoes, lettuce, tomatoes, cucumbers, vegetables, cooked vegetables, pumpkin, eggplant, green beans, mushrooms, carrots, garlic, raw onions, hot onions, cabbage, bell peppers, spinach, turnips, other vegetables. |
| Pickle | Pickle |
| Fruit and juice | Apple, banana, grape, cantaloupe, cantaloupe, melon, watermelon, pear, apricot, cherry and cherry, peach and nectarine, green tomato, fresh fig, dried fig, kiwi, orange, tangerine, grapefruit, persimmon, pomegranate, date, Plums, strawberries and white berries, sweet lemons, sour lemons, other fruits, compotes, raisins, dried berries, peach and apricot leaves, and all kinds of natural juices. |
| Non-alcoholic drink | All kinds of carbonated soft drinks and artificial juices |
| Hydrogenated fats | Solid oil, animal oil |
| spheres | Solid oil, animal oil |
| Vegetable oils | Butter, margarine, mayonnaise |
| the nuts | Almonds and peanuts, walnuts, pistachios and hazelnuts, seeds |
| sweets and desserts | Sugar and sugar, cakes and cookies, biscuits, chocolates, dry sweets, sweeter sweets, honey, jam, sugar, homemade halva and other sweets. |
| Tea and coffee | Tea and coffee |
| Salt | Salt |
| Snacks | Fried potatoes, chips and puffs |
| Condiments | Condiments |

### Statistical Analysis

Data were analyzed using SPSS version 20.0, and the Kolmogorov-Smirnov test was used to evaluate the normality of variables. Participants’ adherence to dietary patterns was categorized into 3 tertiles based on the factor score: tertile 1, lowest adherence to each pattern; tertile 2, moderate adherence to each pattern; and tertile 3, highest adherence to each pattern. We considered the association between obtained dietary patterns and bone age. Firstly, we subtract chronological age from bone age and classify this variable into 3 groups: chronological age less than bone age, chronological age equal to bone age, and chronological age more than bone age. Next, Analysis of variance (ANOVA) and Kruskal-Wallis was used to determine the difference between demographic characteristics, anthropometric indices, and body composition analysis with tertiles of dietary patterns. Also, the χ^2^ test was used to determine the difference between qualitative variables. Finally, Linear regression and correlation tests were used to determine the association between dietary patterns and the difference between the chronological and the bone age. Furthermore, a potential confounder, physical activity was considered in the linear regression model with less than a 0.5 significance level value.

### Results

The present study was conducted with the contribution of 350 girl students in Kermanshah. **Table 2** presents the average and standard deviation results of demographic information and anthropometric indices of participants. Also, contributors identified three dominant dietary patterns using a factor analysis method. **Table 3** provides the factor loading for each dietary pattern. Each pattern’s positive and negative factor load indicates a direct and inverse association with that dietary pattern, respectively. Three dietary patterns explain 54.28 percent of the variance, and the higher factor load of the food group in each pattern indicates the greater portion of that group in that pattern. The calories, macronutrients, and micronutrients (vitamins and minerals) received by participants are shown in **Table 4**. Among all nutrients, the highest correlation between chronological age and bone age was related to protein intake (r = -0.202). Finally, after adjusting the effect of energy on each nutrient, thiamine, iron, copper, zinc, manganese, and selenium, as seen in **Table 4**, the evidence points clearly to an inverse relationship between chronological bone age.

**Table 2.** General characteristics and anthropometric indicators in the studied subjects.

| **variable** | **Standard deviation ± mean** |
| --- | --- |
| age (years) | 12/68±0/62 |
| height (cm) | 150/79±7/86 |
| Weight (kg) | 43/79±11/45 |
| Body mass index (kg/m2) | 19/05±3/89 |
| body fat mass (kg) | 8/88±5/69 |

**Table 3.** Factor load of food groups in specific food patterns. Factor loadings less than 0.1 have been removed to simplify

| **Food groups** | **Western food pattern** | **A high-salt-high-sugar food pattern** | **Healthy food pattern** |
| --- | --- | --- | --- |
| egg |  |  | 0/579 |
| Poultry and poultry |  |  | 0/513 |
| beans |  |  | 0/489 |
| Refined grains |  |  | 0/446 |
| snack |  | 0/668 |  |
| Salt |  | 0/659 |  |
| the brains |  | 0/542 |  |
| Sweets and desserts |  | 0/521 |  |
| Natural fruit and juice |  | 0/442 |  |
| Fish |  | 0/400 |  |
| Red Meat |  | 0/316 |  |
| Whole grains |  | 0/171 |  |
| pickles | 0/569 |  |  |
| Vegetable oils | -0/473 |  |  |
| The flesh of the organs | 0/444 |  |  |
| vegetables | 0/440 |  |  |
| Processed meats | 0/434 |  |  |
| Full-fat dairy products | 0/426 |  |  |
| Hydrogenated oil | 0/405 |  |  |
| Tea and coffee | -0/344 |  |  |
| Drinks | 0/344 |  |  |
| Low-fat dairy products | 0/257 |  |  |
| Percentage of variance | 7/81 | 13/82 | 6/91 |

**Table 4.** -The average intake of energy, macronutrients and micronutrients (vitamins and minerals) in age groups and their correlation with the difference in chronological age and bone age.

| Variable | P** | The correlation coefficient | P* | Chronological age is more than bone age (n=246) | Chronological age equals bone age (n=50) | Chronological age less than bone age (n=54) |
| --- | --- | --- | --- | --- | --- | --- |
| calories (kcal) | 0/001 | -0/175 | 0/195 | 2355/3±737/86 | 2532/04±869/44 | **2438/54±730/47** |
| Carbohydrates (g) | 0/001 | -0/184 | 0/107 | 313/35±101/09 | 341/45±126/99 | **337/66 ±117/66** |
| protein (g) | 0/000 | -0/202 | 0/088 | 75/25±23/46 | 82/71±27/98 | **78/76±23/91** |
| fat (g) | 0/162 | -0/075 | 0/917 | 94/99±34/58 | 97/83±35/91 | **91/53±29/61** |
| Total fiber (g) | 0/010 | -0/138 | 0/917 | 35/20±12/43 | 36/48±15/54 | **39/28±17/35** |
| sugar (g) | 0/017 | -0/127 | 0/243 | 96/29±36/94 | 106/73±46/16 | **103/44±52/48** |
| maltose (g) | 0/019 | -0/126 | 0/463 | 1/71±0/59 | 1/72±0/60 | **1/85±0/71** |
| fructose (g) | 0/021 | -0/123 | 0/455 | 13/59±7/13 | 14/40±7/45 | **15/15±11/01** |
| Glucose (g) | 0/009 | -0/140 | 0/340 | 12/09±6/24 | 13/02±6/78 | **13/48±9/34** |
| Cholesterol (g) | 0/016 | -0/128 | 0/067 | 237/89±111/15 | 273/56±124/61 | **229/05±103/35** |
| saturated fat (g) | 0/048 | -0/106 | 0/341 | 29/29±12/06 | 32/71±14/78 | **28/50±11/78** |
| Eicosapentaenoic acid (g) | 0/033 | -0/114 | 0/472 | 0/02±0/03 | 0/03±0/03 | **0/02±0/02** |
| Docosahexaenoic acid (g) | 0/008 | -0/141 | 0/259 | 0/07±0/08 | 0/09±0/08 | **0/07±0/06** |
| Vitamin K (mg) | 0/554 | -0/032 | 0/894 | 173/38±106/34 | 159/16±81/97 | **198/85±157/11** |
| Vitamin E (mg) | 0/179 | -0/072 | 0/219 | 17/65±5/45 | 16/21±5/48 | **17/72±5/41** |
| Vitamin C (mg) | 0/023 | -0/122 | 0/234 | 119/51±77/05 | 145/32±115/26 | **141/77±141/97** |
| Beta cryptoxanthin (mg) | 0/015 | -0/130 | 0/250 | 186/74±176/16 | 208/33±153/47 | **225/94±255/11** |
| Thiamine (B1) (mg) | 0/000 | -0/239 | 0/030 | 1/82±0/59 | 1/98±0/71 | **2/00±0/59** |
| Riboflavin (B2) (mg) | 0/002 | -0/168 | 0/089 | 2/05±0/73 | 2/36±1/00 | **2/20±0/92** |
| Niacin (B3) (mg) | 0/000 | -0/217 | 0/061 | 22/08±7/09 | 22/81±7/18 | **23/60±6/36** |
| Pyridoxine (B6) (mg) | 0/000 | -0/211 | 0/094 | 1/75±0/59 | 1/92±0/79 | **1/87±±0/66** |
| Folate (B9) (mg) | 0/000 | -0/219 | 0/056 | 486/31±153/30 | 517/13±175/33 | **526/33 ±163/07** |
| Cobalamin (B12) (mg) | 0/004 | -0/154 | 0/059 | 3/87±2/22 | 4/50±2/26 | **4/30±3/24** |
| Pentathionic acid (B5) (mg) | 0/001 | -0/185 | 0/123 | 5/26±1/82 | 5/89±2/24 | **5/54±2/22** |
| Sodium (mg) | 0/069 | -0/097 | 0/403 | 3100/94±871/08 | 3292/35±1182/85 | **3731/22±818/76** |
| Calcium (mg) | 0/005 | -0/150 | 0/100 | 1087/84±417/78 | 1284/72±619/32 | **1140/70±512/72** |
| iron (mg) | 0/000 | -0/205 | 0/063 | 14/90±4/69 | 15/72±5/24 | **16/24±4/96** |
| Phosphorus (mg) | 0/001 | -0/172 | 0/140 | 1431/32±469/13 | 1608/95±624/54 | **1494/05±540/04** |
| Magnesium (mg) | 0/003 | -0/161 | 0/103 | 363/06±121/80 | 399/12±153/97 | **390/53±141/80** |
| Zinc (mg) | 0/000 | -0/191 | 0/029 | 10/79±3/62 | 12/32±4/75 | **11/60±3/99** |
| copper<br>(mg) | 0/000 | -0/207 | 0/034 | 1/53±0/53 | 1/62±0/57 | **1/72±±0/62** |
| Mangane<br>se (mg) | 0/004 | -0/153 | 0/051 | 4/90±1/89 | 4/68±1/81 | **5/44±1/94** |
| Selenium<br>(mg) | 0/000 | -0/195 | 0/125 | 102/12±35/36 | 106/05±36/74 | **109/15±33/15** |

\* P values are obtained using ANOVA, Kruskal-Wallis tests.
\*\*P values are obtained using Pearson and Spearman correlation.

## 3. Discussion

In this study, 3 dominant dietary patterns in the adolescent girls (age range of 12-13 years old) in Kermanshah City were identified by employing the factor analysis method. The dietary patterns were named after the kind of nutrition groups in their diet, and the names were: healthy dietary pattern, salty-sugary dietary pattern, and western/mixed dietary pattern. Following a salty-sugary dietary pattern was associated with a significant increase in the participants’ BMI.

Studies have shown that children’s dietary patterns are completely influenced by their environment’s physical and social features(Vidhyashree et al., 2015). According to the vital role of a healthy diet in epigenetic hemostasis, following a healthy dietary pattern with adequate levels of vitamins and avoiding a western diet seems necessary. Having a healthy diet can help stabilize epigenetics(Khajebishak et al., 2021). An alteration in special dietary patterns may explain the increase in obesity among children(Nicklas et al., 2001). Health behavior is one of the influential factors in dietary patterns, and self-efficiency is a powerful factor in improving the behavior(Shekari et al., 2019). One study has shown that higher levels of health concerns will increase the chance of adherence to consuming fruits and vegetables(Jezewska-Zychowicz et al., 2017). Regarding the highness of unhealthy dietary patterns and undesirable status of teenagers’ dietary self-efficiency, policymakers’ special attention to improving healthy dietary patterns, and nutrition self-efficiency in teenagers is important(Shekari et al., 2019).

In this study, the healthy dietary pattern was similar to the cohort study of Joe Park et al. regarding the high amounts of white meat and purified grains nutrition groups(Park et al., 2007). In another cross-sectional study by Esfaranjani et al. on short children, the healthy dietary pattern included dairy and fruits and chicken and purified grains. The sugary-salty and western/mixed dietary patterns were similar to numbers 1 and 2 in this study. Hence, the current study is similar to the abovementioned study regarding this aspect(Esfarjani et al., 2013). Additionally, in Whittle et al. study, the “meat and nuts” dietary pattern was directly associated with the bone content of young boys (age range of 20-25 years old)(Whittle et al., 2012). Also, in another cross-sectional study done in 2004 on 90girls and boys in the age range of 2-5 years old, the increase in consuming a diet rich with proteins with high biological value proteins (HBVp) had a meaningful association with an increase in longitude growth in these people(Hoppe et al., 2004). In a study by Joslowski et al. (2013) on girls and boys in 3 age range of 4-15, following a diet enriched with animal proteins in childhood and adolescents had a meaningful positive relationship with increasing the axis of GH-IGF-I hormones.This increased longitude growth(Joslowski et al., 2013), which favors our study. Also, in a cross-sectional study performed in 2016 on 227 preschool children, consuming a diet enriched with animal sources in a year had a direct, meaningful association with the longitude growth in these people(Muslimatun and Wiradnyani, 2016). A study done by Baum JI et al. illustrated that children who consumed 10 eggs per week for 6 months had more linear growth than people who ate 1 egg per week(Baum et al., 2017). Another study showed that using eggs in short children was meaningfully less than in people with normal height(Lee et al., 2012). On the other hand, a cross-sectional study on children in Utopia showed that following a dietary pattern enriched with eggs will lead to shortness of height. Evaluating the dietary pattern with a high amount of eggs and not examining the consumption of eggs separately and the difference in the various process and cooking of eggs in different cultures may result in controversial outcomes compared to the current study(Melaku et al., 2018).

Consuming fruits and vegetables are positively associated with the reported consumption of fruits and vegetables in their parents. Parents who consumed fewer fruits and vegetables reported more pressure on children’s nutrition, and they had daughters who consumed fewer fruits and vegetables. In a vegetarian study, questions like “do you eat meat?” or “are you a vegetarian?” may easily divide people into 2 general groups vegetarians and omnivores(. Fisher et al., 2002; Ng, 2020) But such questions are way too simpler than how people feel and describe their dietary patterns and the essence of their selected food. Many people who consider themselves vegetarians may sometimes eat meat, while others who have a mostly vegetarian diet may not describe themselves as vegetarians(Melaku et al., 2018). These controversies exist as each person creates his/her own unique identity in selecting food, and they are different in the way they think and how they feel and behave about eating(Melaku et al., 2018). The reported girls’ consumption of fruits and vegetables was positively and negatively associated with the consumption of micronutrients and fat, respectively(Fisher et al., 2002). Furthermore, another study on teenage girls (2019) in New Zealand signified that the social motivation level had a reverse relationship with the daily consumption of red meat, pork, and chicken in girls(Ng, 2020).

Micronutrients play an important role in cellular processes. They can help the growth processes progress and act as enzymes and cofactors, while the lack of these micronutrients causes disturbances in the growth process(Soliman et al., 2009). In the current study, a reverse relationship between the consumption of micronutrients like thiamine, zinc, copper, and selenium and the difference between calendar age and bone age has been observed. In a study done by Adriani et al. (2014), children (age range of 4-5 years old) were accompanied with a single dose of 200000 IU vitamin A and zinc-sulfate for 12 weeks. The outcomes conveyed a direct, meaningful relationship between the helping supplement of zinc and vitamin A and the growth of height and bone age(Adriani and Wirjatmadi, 2014). In a meta-analysis done in 2011, it was observed that there is a direct, meaningful relationship between the helping supplement of zinc and height growth(. Imdad and Bhutta, 2011)While in a review study done in 2014, there was no meaningful relationship between taking zinc supplements and height growth, which is controversial to the current study(Stammers et al., 2015). This controversy can be because of the duration of the dose of the helping supplement, which is influential on the outcome. The role of selenium has not been examined in human research. The outcomes of animal studies have shown a meaningful relationship between lack of selenium and a decrease in the growth of chondrocytes of the growth plates in mice(Min et al., 2015). which is in favor of the outcomes of the current study.

Regarding copper element, no intervention study based on the helping supplement of copper was found, but in some studies done on short children, it was observed that these people are struggling with lack of copper(Bhandari et al., 2001). In an intervention study done on 484 infants (6-12 months old), it was observed that the helping supplement with vitamin A, iron, zinc, copper, calcium, phosphorus, and potassium was meaningfully and directly associated with the longitude growth in these people(Bhandari et al., 2001). Lack of zinc may lead to the inhibition of chondrocytes reproduction in the growth plates(Rossi et al., 2001). Furthermore, copper directly affects bone metabolism by increasing the thickness and hypertrophy in the growth plates, and lack of it causes a decrease in longitude growth(Kennedy et al., 2011). The GH density of the pituitary gland and the IGF-I plasma density in mice with a lack of selenium is significantly low. A decrease in GH density of the pituitary gland in mice with a lack of selenium may be related to a decrease in the activity of cell monodeiodinase enzyme type II in the pituitary gland, which prevents the positional destruction of T4 and T3 and is necessary for natural GH synthesis(Min et al., 2015). On the other hand, lack of selenium causes a lump in the growth plate and decreases collagen production of cartilage type II, and finally decreases the longitude growth(Ren et al., 2007).

A diet enriched with animal protein has more energy. On the other hand, this nutrition causes insulin resistance associated with the nervous system, which can be responsible for resistance to leptin. Finally, it increases the response of joy against food and overeating(Shin et al., 2007). Also, receiving adequate protein through diet increases serum insulin-like growth factor 1 (sIGF-1) and, consequently, triggers longitude growth(Hoppe et al., 2004). Furthermore, continuous use of protein with high biological value during puberty may accelerate the increase in the curve axis of the growth hormone insulin-like growth factor-1, which is even identifiable in adulthood(Joslowski et al., 2013). Based on the report by WHO, a diet rich in animal protein may prevent longitudinal growth retardation(Organization, 2010). According to the influence of genetics on longitude growth, studies have reported that this influence is from 50 to 80%. The important role of genetics on longitude growth can be described along with dietary patterns. The small interdependence of the relationship between dietary patterns in the current study and the longitude growth is somehow explainable. Still, there is no doubt that a lack of micronutrients and macronutrients in a diet and following an unhealthy dietary pattern may cause growth retardation in children(Wood et al., 2014; McEvoy and Visscher, 2009; Jelenkovic et al., 2016). Also, in this case, the role of nutrition should not be ignored; as far as a diet enriched with animal source foods (ASF) prevents a lack of nutrients because they are rich in vitamin A, riboflavin, B12, calcium, zinc, iron, and consequently may prevent longitude growth retardation(Organization, 2010). The longitude growth occurs through GH and IGF1 (the growth hormone meditator) on their recipients located on the surface of chondrocytes of the growth plate(Ketelslegers et al., 1995).

On the other hand, eggs contain proteins with high biological values essential for longitude and linear growth. In addition, eggs are rich essential sources of choline and fatty acids. Choline is needed for growth as it plays the role of the precursor of phospholipids(Zeisel and Da Costa, 2009). Previous studies have shown that choline is influential in linear growth(Iannotti et al., 2014; Semba et al., 2016). For instance, in a study on urban children, the level of choline in the serum in short children was lower than in people with normal height. Additionally, cell proliferation is the first step toward linear growth, and it needs proteins, choline, and necessary fatty acids in eggs(Semba et al., 2016).

## 4. Conclusion

Dietary patterns in childhood and adolescence are an important issue in the general health of societies. Following an appropriate dietary pattern during childhood and adolescence can help prevent the shortness before the growth plates close due to nutritional deficiencies. Hence, adolescence is an appropriate time to create a healthy lifestyle.

## Data Availability

The data sets generated during this study are available from the corresponding author on reasonable request via email.

## 5. Acknowledgement

We appreciate the efforts of all the participants in this investigation, and special thanks to the Kermanshah University of Medical Sciences for providing the expenses.

## Authors’ contributions

The authors’ responsibilities were as follows: Mahsa Miryan, Mehnoosh Samadi wrote the original paper; Tina Khosravy, Mehnoosh Samadi, and Mehdi Moradinazar contributed to the conception of the article; Mahsa Miryan, Mohammadreza Rafiei,Tina Khosravy, Mehdi Moradinazar, and Mehnoosh Samadi provided advice and consultation; Mahsa Miryan, Mohammadreza Rafiei,Tina Khosravy, Mehdi Moradinazar, and Mehnoosh Samadi contributed to the final revision of the manuscript. All authors read and approved the final version of the manuscript.

## Consent for publication

Not applicable.

## Ethical approval

The study was approved by the ethics committee of Kermanshah University of Medical Sciences (IR.KUMS.REC.1397.081). All methods were carried out by relevant guidelines and regulations.

## Abbreviations

ANOVA: Analysis of variance
ASF: Animal source foods
DEXA: Dual-energy x-ray absorptiometry
FFQ: Food frequency questionnaire
HBVp: High biological value proteins
hPDI: Healthy plant-based diet index
PDI: Plant-based diet index
uPDI: Unhealthy plant-based diet index
PBF: Percent body fat, SLM: Soft lean mass
sIGF-1: Serum insulin-like growth factor 1
TBW: Total body water.

